# Health coaching 2.0: redefining a core lifestyle medicine intervention through a structured, BCT-based model

**DOI:** 10.1101/2024.06.10.24308710

**Authors:** Ari Langinkoski, Johanna Hämäläinen, Krista Rapo, Mikko Rinta, Pekka Aroviita, Mika Venojärvi

**Affiliations:** School of Medicine, Institute of Biomedicine: Sports and Exercise Medicine, University of Eastern Finland, Kuopio, Finland; Nordic Health Academy, Helsinki, Finland; Terveystalo Oyj, Helsinki, Finland

**Keywords:** health coaching, behavior change, lifestyle medicine, digital health, preventive care

## Abstract

**Background:** Preventing non-communicable diseases (NCDs) is a global health priority, as they remain the leading cause of mortality globally, responsible for over 40 million deaths each year. Health coaching has demonstrated potential to improve lifestyle behaviors and health outcomes, yet its practical application is limited by inconsistent definitions and training standards.

**Objective:** This brief report evaluates the feasibility and preliminary effectiveness of a systematically defined health coaching model—PrioMed®—that translates the coaching process into evidence-based Behavior Change Techniques (BCTs), providing a structured and teachable framework for healthcare delivery.

**Methods:** A single-arm pilot study (n = 25) was conducted in private healthcare in Finland. Two registered nurses without prior coaching experience were trained for three half-day sessions and used a 25-page manual to deliver PrioMed® coaching across seven phone-based sessions over six months. Participants completed baseline and follow-up surveys, including the PROMIS-10 Global Health instrument. Paired-sample t tests and Cohen’s d were used to assess changes in physical and mental health T-scores.

**Results:** Participants (mean age 61.9 years, 76% female) showed significant improvements in self-reported health. Physical health increased by +5.72 T-points (p < .001, d = 1.31) and mental health by +4.43 T-points (p < .001, d = 0.83), both indicating large effect sizes.

**Conclusions:** Redefining health coaching through an explicit BCT framework appears feasible, teachable, and capable of generating meaningful health improvements within limited coaching time. The PrioMed® model offers a replicable approach for digital and nurse-led lifestyle interventions that warrants further evaluation in controlled trials and more diverse real-world populations.

## Introduction

Non-communicable diseases (NCDs) remain the leading cause of global mortality, accounting for approximately 43 million deaths annually and 75% of non-pandemic deaths worldwide (1). The majority of these conditions—such as cardiovascular diseases, type 2 diabetes, chronic respiratory diseases, and certain cancers—are strongly associated with modifiable lifestyle factors including diet, physical inactivity, alcohol use, smoking, and stress-related behaviors (2). Addressing these determinants through effective, scalable, and sustainable interventions remains a cornerstone of preventive health and lifestyle medicine.

Health coaching has emerged as one of the most promising methods for facilitating sustained lifestyle change in both preventive and clinical contexts. Systematic reviews and meta-analyses (3–5) indicate that health coaching can enhance self-efficacy, improve health behaviors, and contribute to better clinical outcomes, often with favorable cost-effectiveness profiles (6). However, despite its growing popularity, *health coaching* remains ambiguously defined across the literature and professional practice. The concept has been variably operationalized—from motivational interviewing and counseling approaches to comprehensive lifestyle medicine interventions—resulting in considerable heterogeneity in both methodology and reported outcomes (4, 5, 7).

This lack of a shared operational definition poses two key challenges. First, it impedes the reproducibility and scientific validation of health coaching interventions. Second, it hinders the ability to train health professionals consistently and efficiently. Without a structured and evidence-based framework, coaching programs often rely on individual experience rather than standardized methods, limiting their scalability within healthcare systems.

To address these challenges, we developed and tested a new coaching model—PrioMed® Health Coaching—that systematically defines the coaching process using the Behavior Change Technique (BCT) taxonomy v1 (8). The model is grounded in the six pillars of lifestyle medicine (9) and integrates only those BCTs with established evidence of effectiveness (10). By translating the traditionally abstract concept of “coaching” into a set of clearly defined, teachable, and measurable behavior change components, the PrioMed® model aims to make the coaching process both scientifically replicable and practically scalable.

In contrast to previous coaching programs that require extensive training, PrioMed® has been specifically engineered for rapid integration into clinical practice. In this pilot, two nurses with no prior coaching experience received concise theoretical and practical training and implemented the intervention with their clients in a private healthcare environment. The training was delivered by the Nordic Health Academy, in collaboration with the University of Eastern Finland and Terveystalo Oyj, a large private healthcare provider. Supplementary data from a related study support the acceptability and feasibility of the PrioMed® training framework, demonstrating that both coaches and clients viewed the integration of the health coaching into routine clinical practice as practical and well-received.

From a digital health perspective, the PrioMed® model is particularly relevant because it represents a structured, data-compatible coaching process that can be readily integrated with digital monitoring tools and remote care pathways. Each phase of the coaching intervention— goal setting, needs analysis, feedback, and review—corresponds to distinct, codifiable BCT elements. This opens the door to future digital augmentation through wearable data, decision-support algorithms, and artificial intelligence–assisted behavior tracking, enabling personalized and continuous lifestyle support at scale.

The present study reports the findings from the initial feasibility and effectiveness evaluation of the PrioMed® pilot program. Conducted in a real-world private healthcare setting, this pilot explored whether health coaching, redefined through a structured BCT-based framework, could produce measurable improvements in self-reported physical and mental health within a limited intervention time. Specifically, the study aimed to (A) assess the feasibility of implementing the PrioMed® model in practice, and (B) evaluate preliminary health outcomes following a six-month intervention delivered by newly trained nurses.

By addressing long-standing issues in the conceptual clarity and standardization of health coaching, the PrioMed® model seeks to provide a universal, evidence-based structure that bridges the gap between lifestyle medicine theory and scalable digital health implementation.

## Methods

### Study design and setting

This single-arm, six-month pilot study evaluated the feasibility and preliminary effectiveness of the PrioMed® Health Coaching model in a real-world private healthcare environment. The study was conducted at Terveystalo Oyj, one of Finland’s largest private healthcare providers, in collaboration with the Nordic Health Academy and the University of Eastern Finland.

The study adhered to the Declaration of Helsinki and the ICMJE recommendations for research involving human participants. Ethical approval for the study was obtained from The Regional Medical Research Ethics Committee of Eastern Finland Collaborative Area and all participants provided written informed consent prior to participation.

### Participants and recruitment

Participants were recruited from Terveystalo’s existing client base, including those who had previously participated in an annual health check-up or a digital wellness program. Inclusion criteria were age ≥18 years, adequate Finnish language skills, and willingness to participate in remote coaching. Exclusion criteria included acute medical conditions or cognitive impairments preventing informed participation.

A total of 25 participants completed both baseline and six-month follow-up assessments. The demographic characteristics are presented in Table 1.

**Table 1.** Participants demographics.

| **Characteristic** | **Value** |
| --- | --- |
| Participants, n | 25 |
| Age, mean ± SD (range) | 61.9 ± 10.8 (33–75) |
| Female, n (%) | 19 (76%) |
| Married or partnered, n (%) | 18 (72%) |
| Retired or not currently working, n (%) | 12 (48%) |
| Post-secondary or higher education, n (%) | 23 (92%) |

### Intervention

The PrioMed® model operationalizes health coaching through evidence-based Behavior Change Techniques (BCTs) drawn from the taxonomy by Michie et al. (8) and informed by the six pillars of Lifestyle Medicine (9). It provides a structured and teachable framework emphasizing self-assessment, needs analysis, goal setting, graded progression, and feedback.

Two registered nurses with no prior coaching experience were trained by the first author in three four-hour sessions using a 25-page instructor manual. The training combined theoretical instruction and practical exercises focusing on motivational communication, goal-oriented dialogue, and structured session management.

Each participant received up to seven phone-based sessions across six months, combining an initial assessment and goal-setting phase with five to six follow-up sessions focused on feedback and reinforcement. Total contact time averaged less than four hours per participant.

A detailed description of the PrioMed® coaching process, including session structure, BCT mapping, and sample tools (PrioMed® Lifestyle Medicine Vital Signs [PMLMVS] and PrioMed® Needs Analysis [PMNA]), is provided in Supplementary Material 1.

### Outcome measures

The primary outcomes were self-reported physical and mental health, measured using the PROMIS-10 Global Health questionnaire, which produces standardized T-scores (mean = 50, SD = 10; higher scores indicate better health). Surveys were completed electronically at baseline and at six-month follow-up.

Secondary outcomes included demographic data and descriptive summaries of lifestyle self-assessments (PMLMVS) and applied BCT frequencies recorded by the coaches.

### Data analysis

PROMIS-10 raw scores were converted to T-scores using the *HealthMeasures Scoring Service*. Data normality was verified with the Shapiro–Wilk test, and pre-post differences were analyzed using paired-sample t-tests. Effect sizes were expressed as Cohen’s d, using the standard deviation of the mean difference as the denominator. The threshold for statistical significance was *p* < 0.05 (two-tailed). Analyses were conducted in IBM SPSS Statistics v29. Complementary qualitative findings related to training and user experience are presented in a companion study (11).

## Results

### Participant characteristics

Twenty-five participants completed both baseline and six-month follow-up surveys. Demographic characteristics are summarized in Table 1. Participants were predominantly older, well-educated adults, mostly women, and nearly half were retired or not currently working.

### Primary outcomes

Statistically significant improvements were observed in both physical and mental health domains of the PROMIS-10 Global Health measure after six months of PrioMed® health coaching.

- Physical Health T-score: Baseline mean = 45.51 ± 5.59; follow-up mean = 51.23 ± 5.04; mean change = + 5.72 points (95 % CI 3.92–7.51; *p* < .001).
- Mental Health T-score: Baseline mean = 50.20 ± 6.91; follow-up mean = 54.63 ± 5.47; mean change = + 4.43 points (95 % CI 2.24–6.62; *p* < .001).

Effect sizes were large for both scales (Cohen’s *d* = 1.31 for physical health; *d* = 0.83 for mental health), suggesting clinically meaningful changes in perceived wellbeing.

### Secondary observations

Analysis of coaching session documentation indicated high adherence to the intended PrioMed® protocol. The most frequently used Behavior Change Techniques (BCTs) were:

- *Goal setting (behavior)* – 100 % of participants
- *Goal setting (outcome)* – 88 %
- *Self-monitoring of behavior* – 68 %
- *Social reward* – 64 %
- *Review behavior goals* – 60 %
- *Comparative imagining of future outcomes* – 60 %
- *Action planning* – 56 %.

Other BCTs were applied in fewer than half of the coaching relationships, reflecting the limited duration and goal-oriented nature of the intervention. Average total contact time remained below four hours per participant, confirming the intended efficiency of the model.

### Feasibility and implementation findings

From an implementation standpoint, the PrioMed® model proved feasible for delivery by nurses with no prior coaching experience. Both coaches successfully completed the training, adhered to the structured protocol, and maintained consistent client engagement throughout the study. No technical or logistical difficulties were reported in the telephone-based format.

Complementary qualitative findings, reported in a companion study (11), reinforce these findings: Coachees characterized the sessions as supportive, personalized, and motivating, while coaches reported marked improvements in their confidence and competence over the six-month period.

## Discussion

This pilot study evaluated the feasibility and preliminary effectiveness of a newly developed health coaching model grounded in BCTs, within a real-world private healthcare setting. Despite the brief duration and limited contact time, the intervention was associated with notable improvements in participants’ self-reported physical and mental health outcomes. These findings indicate that a structured, behaviorally defined coaching framework can feasibly be implemented and may contribute to enhanced subjective wellbeing, even when delivered with a minimal resource investment.

### Interpretation of main findings

The observed effect sizes in the present study are substantially larger than those typically reported in lifestyle intervention meta-analyses (12, 13). From clinical perspective, changes in this magnitude are typically regarded as meaningful, reflecting noticeable enhancement in functional capacity and reduction symptom burden. The observed gains are particularly notable considering the brevity of the intervention (< 4 h per participant) and the fact that coaches were newly trained nurses with no prior experience in behavioral counseling. Together, these results suggest that redefining health coaching through clearly operationalized BCTs can enhance both its efficiency and trainability.

Documentation from the coaching sessions demonstrated high protocol adherence, with frequent use of core BCTs such as goal setting, self-monitoring, and action planning. This supports the fidelity of the intervention and reinforces its theoretical grounding.

### Comparison with previous literature

Previous systematic reviews have identified health coaching as a potentially effective yet highly heterogeneous intervention domain (3, 4, 5). Variability in definitions, training, and delivery formats has hindered replication and large-scale implementation. By explicitly structuring the PrioMed® model around empirically supported BCTs (8, 10), this study addresses a long-standing gap in the literature: the need for a reproducible, evidence-linked framework that preserves the person-centered ethos of coaching while enabling systematic evaluation.

The present results are consistent with emerging evidence showing that targeted, time-limited coaching interventions can yield measurable health benefits, particularly when they include goal setting, self-monitoring (12–14), and graded tasks (13). The structured yet autonomy-supportive design of PrioMed® may explain the strong adherence observed among participants. Additional qualitative insights from the companion study (11) further supports the feasibility of intervention. Both coaches and coachees reported high levels of satisfaction and perceived the sessions as relevant and beneficial, even delivered via remote modalities (11).

### Practical implications and digital health relevance

From an implementation perspective, these findings support the integration of structured health coaching models into both primary care and digital health ecosystems. The PrioMed® framework—by defining the coaching process in modular, codifiable BCT units—creates a foundation for digitally augmented coaching, in which wearable-derived data and algorithmic decision support can inform personalized behavioral feedback loops. Such standardization also enables quality assurance and competence-based training for health professionals, addressing a key barrier to scaling lifestyle interventions in healthcare systems.

In practical terms, the model demonstrates that brief, remote coaching interactions can be sufficient to produce meaningful improvements in perceived health status. This efficiency is particularly relevant for preventive care models that seek to reach large populations at moderate cost. The structured PrioMed® model further allows delegation of coaching to non-specialist healthcare staff, such as nurses or health advisors, thereby expanding access while maintaining methodological fidelity.

Within the broader digital health landscape, the PrioMed® approach aligns with current priorities in data-driven, personalized preventive healthcare—an area increasingly supported by wearable sensors, continuous monitoring, and AI-enabled behavior analytics. The BCT-based logic of the model provides an interpretable bridge between behavioral science and digital system design, facilitating future research on how human and automated coaching elements can complement each other.

### Strengths, limitations, and future directions

The main strengths of this pilot include its pragmatic design, use of a validated outcome measure (PROMIS-10), and detailed documentation of coaching content using BCT taxonomy. Conducting the study within an active private healthcare setting enhances ecological validity and demonstrates feasibility under real-world conditions.

However, some limitations must be acknowledged. The study lacked a control group and randomization, limiting causal inference. The relatively small, self-enrolled sample— predominantly older, well-educated women—restricts generalizability. Furthermore, the coaches’ dual roles as healthcare employees may have introduced unmeasured contextual effects. Outcomes were self-reported, and objective behavioral or physiological measures (e.g., activity tracking, biometrics) were not included.

Future studies should therefore employ randomized controlled designs, larger and more diverse samples, and multimodal outcome measures integrating both subjective and digital health data. Longer-term follow-up would also clarify the sustainability of observed changes. Additional research could evaluate the PrioMed® framework’s integration with wearable technologies and adaptive algorithms, as well as its cost-effectiveness in large-scale preventive health programs.

## Conclusion

Overall, the pilot demonstrated that a standardized, BCT-based coaching model can be effectively implemented in private healthcare, achieving meaningful improvements in health-related quality of life with minimal time investment. The findings support continued development of standardized and scalable coaching frameworks that can effectively integrate principles of lifestyle medicine with emerging innovations in digital health.

## Data Availability

All data produced in the present study are available upon reasonable request to the authors

## Declarations

### Ethics approval and consent to participate

The study was approved by The Regional Medical Research Ethics Committee of Eastern Finland Collaborative Area (approval number: [1121/13.02.00/2022]). All participants provided written informed consent prior to enrollment, and the study was conducted in accordance with the principles of the Declaration of Helsinki.

### Consent for publication

Not applicable.

### Author contributions

- **Ari Langinkoski:** Conceptualization, methodology, training design, data analysis, writing – original draft.
- **Johanna Hämäläinen:** Data collection, survey analysis, writing – review & editing.
- **Krista Rapo:** Data collection, survey analysis, writing – review & editing.
- **Mikko Rinta:** Methodological input, nutrition expertise, writing – review & editing.
- **Pekka Aroviita:** Medical oversight, study supervision, writing – review & editing.
- **Mika Venojärvi:** Supervision, methodology, writing – review & editing.

All authors have read and approved the final manuscript and agree to be accountable for all aspects of the work.

## Acknowledgements

The authors thank the participating nurses and study participants for their valuable contributions. The authors also acknowledge the use of OpenAI’s ChatGPT (GPT-5, 2025) to assist in language editing, reference formatting, and structural alignment with the *Digital Health* author guidelines. The authors remain fully responsible for the content and accuracy of this manuscript.

## Availability of data and materials

Deidentified participant data are available from the corresponding author upon reasonable request. Supplementary materials describing the intervention are available as Supplementary Material 1.

