## Supplementary Material 1 for "Health coaching 2.0: redefining a core lifestyle medicine intervention through a structured, BCT-based model"

### ***PrioMed® Coaching Process***

PrioMed® Health and Lifestyle coaching uses a systemized method (Figure 1) based on BCTs that have evidence of their effectiveness and is centered around goal setting and graded steps. To emphasize the autonomy supportive and patient centered method the coaching process begins with a self-evaluation of the six pillars of lifestyle medicine (PrioMed® Lifestyle Medicine Vital Signs [PMLMVS]). After reflection with the coach, the patient self-selects the lifestyle he or she wishes to focus on.

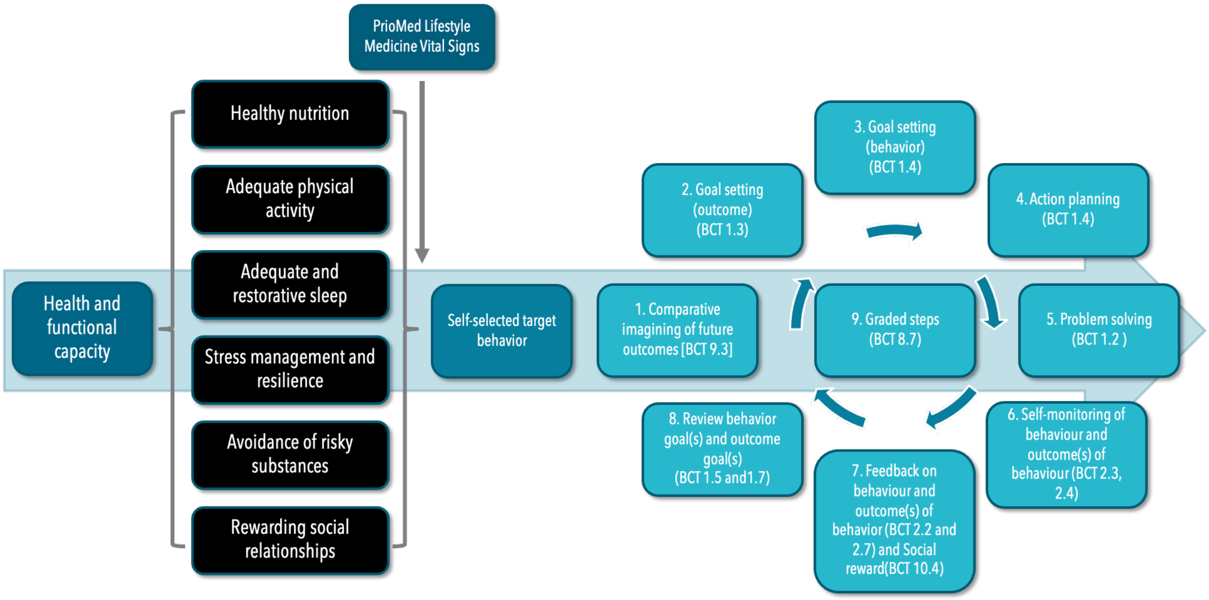


**Figure 1**. PrioMed® Lifestyle Medicine Vital Signs and PrioMed® Goal Setting Wheel. Behavior Change Techniques (BCT) from Michie et al. (2013).

#### *The Structure of the coaching sessions*

The structure of the coaching sessions is presented in Table 1. The actual number of sessions ranged from 4–7 ($M$ = 6.04 $SD$ = 0.84) and total time spend was <4 hours per participant.

**Table 1**. Structure and content of coaching sessions

| Session | Duration | Main components |
|---|---|---|
| **1** | 45 min | Reflecting on PMLMVS; selecting the lifestyle domain of focus; initial goal setting (if time allowed) |
| **2** | 45 min | Reflecting on PrioMed® Needs Analysis (PMNA); finalizing the main goal |
| **3** | 15 min | Feedback and social reward (if appropriate) |
| **4** | 15 min | Feedback and social reward (if appropriate) |
| **5** | 30 min | Feedback and social reward (if appropriate); reviewing goals |
| **6** | 15 min | Feedback and social reward (if appropriate) |
| **7** | 15 min (+30 min if surveys not completed earlier) | Feedback and social reward (if appropriate); collection of follow-up surveys |

#### *PrioMed® Needs Analysis*

The PMNA was inspired by a process that is typical, if not an integral part, of elite sports coaching. To our knowledge, this is not well documented in peer-reviewed literature, but the basic idea is this: First, you analyze the demands of the sport, e.g., strength, speed, agility & stamina. Then, you analyze the athlete on the same characteristics. Finally, you do a comparative analysis between the sports demands and the athlete, and the result is a precise needs analysis of what you need to focus on in the athlete's training.

In the PMNA the "sport" is the lifestyle in focus, and the demands were taken from the COM-B model (Michie et al., 2011), which was specifically developed to serve as a logical foundation for interventions.

The participants completed the PMNA self-evaluation between the first and the second meeting. After a reflection whether some determinant might need special attention, the coach was encouraged to use the following BCTs (Figure 2), after which the process continued as per Figure 1. The exact definitions of each BCT can be found on the electronic supplementary material of BCT taxonomy v1 (Michie et al., 2013).

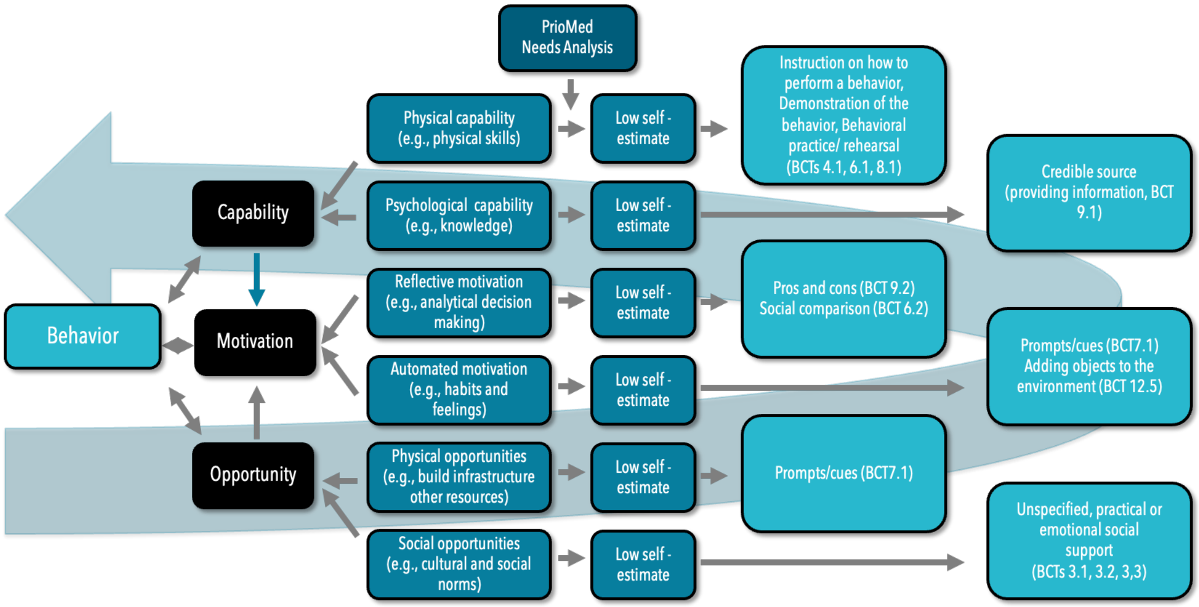


**Figure 2.** PrioMed® Needs Analysis. Behavior Change Techniques (BCT) from Michie et al. (2013).

**PrioMed® Lifestyle Medicine Vital Signs Questionnaire**

(NOTE: The questionnaires used in the study were in Finnish language, here an English translation is presented.)

Please evaluate every statement on a scale of 0–10, where 0 means *I strongly disagree* and 10 means *I strongly agree*.

| | | | | | | | | | | |
|---|---|---|---|---|---|---|---|---|---|---|
| I strongly | | | | | | | | | | I strongly |
| disagree | | | | | | | | | | Agree |
| 0 | 1 | 2 | 3 | 4 | 5 | 6 | 7 | 8 | 9 | 10 |
| ☐ | ☐ | ☐ | ☐ | ☐ | ☐ | ☐ | ☐ | ☐ | ☐ | ☐ |

1. In my perception, my eating habits promote my well-being and health.

2. In my perception, my physical activity levels are adequate for promoting my well-being and health.

3. In my perception, I get adequate amounts of good quality sleep to promote my well-being and health.

4. In my perception, my mental strain (feeling of stress) is on a level that does not hinder my well-being or health.

5. In my perception, if or when I use substances (such as alcohol or tobacco), my use is on a level that does not hinder my well-being or health.

6. In my perception, my social relationships are rewarding and promote my well-being and health.

(Perception = the way in which something is regarded, understood, or interpreted. Almost the same as I believe...)

**Example of PrioMed® Needs Analysis Questionnaire (Nutrition)**

(NOTE: The questionnaires used in the study were in Finnish language, here an English translation is presented.)

Please evaluate every statement on a scale of 0–10, where 0 means *I strongly disagree* and 10 means *I strongly agree*.

| | | | | | | | | | | |
|---|---|---|---|---|---|---|---|---|---|---|
| I strongly | | | | | | | | | | I strongly |
| disagree | | | | | | | | | | Agree |
| 0 | 1 | 2 | 3 | 4 | 5 | 6 | 7 | 8 | 9 | 10 |
| ☐ | ☐ | ☐ | ☐ | ☐ | ☐ | ☐ | ☐ | ☐ | ☐ | ☐ |

**NUTRITION**

1. I know well which eating habits best promote my health. In addition, I possess the practical skills necessary to prepare healthy meals and maintain eating habits that enhance my well-being.

2. I know how to make healthier nutritional choices, and I have the resources to maintain these healthier choices even in challenging conditions and situations.

3. I am well-informed about the health effects of nutrition. I have contemplated the importance of nutrition and its impact on my health and well-being. I am also motivated to find ways to eat more healthily.

4. I have established behaviors or routines that I naturally follow in my daily life, which help me make healthy nutrition choices.

5. My life situation and physical environment enable the consumption of healthy food. For instance, I have time to prepare healthy meals, and healthy options are available where I usually eat or purchase my food.

6. My social environment, such as family members, friends, or colleagues, values the importance of healthy nutrition and supports my efforts to eat more healthily.

***Supplementary Material References***

Michie S, van Stralen MM, West R. The behaviour change wheel: a new method for characterising and designing behaviour change interventions. Implement Sci. 2011;6:42. doi:10.1186/1748-5908-6-42.

Michie S, Richardson M, Johnston M, et al. The behavior change technique taxonomy (v1) of 93 hierarchically clustered techniques: building an international consensus for the reporting of behavior change interventions. Ann Behav Med 2013;46:81–95. doi:10.1007/s12160-013-9486-6
